# Ultrasound-Guided Rete Testis Approach to Sperm Aspiration and Spermatogonial Stem Cell Transplantation in Patients with Azoospermia

**DOI:** 10.1101/2025.03.25.25324518

**Authors:** Amanda Colvin Zielen, Karen A. Peters, Gunapala Shetty, Deborah A. Gross, Carol B. Hanna, Serena L. Dovey, Anna Wecht, Glenn M. Cannon, Marvin L. Meistrich, Michael Hsieh, Kathleen Hwang, Kyle E. Orwig

## Abstract

**Background:** Azoospermia, characterized by the absence of sperm in the ejaculate, impacts 1% of men globally. Surgical approaches to retrieve sperm from the testis are effective in some cases but can be invasive and expensive. Here we described a noninvasive ultrasound-guided rete testis (UGRT) approach to retrieve sperm from the testes or infuse stem cells into the testes of men with azoospermia.

**Methods:** Ultrasound was used to guide insertion of a 25-gauge hypodermic needle through the base of the scrotum and into the rete testis space that is contiguous with all seminiferous tubules. Medium was infused into the rete testis and aspirated to retrieve sperm from monkeys and men (NCT03291522) with azoospermia. A peripubertal patient with osteosarcoma of the femur cryopreserved a testicular cell suspension to safeguard his future fertility (NCT02972801). He returned as a young adult (21-25 yo) survivor for autologous transplantation of his cryopreserved testicular cells, including spermatogonial stem cells, using the UGRT approach (NCT04452305).

**Findings:** Sperm were successfully aspirated from four of six monkeys with radiation-induced azoospermia and three of three patients with obstructive azoospermia, demonstrating proficiency with the UGRT approach. Sperm were not recovered from the testes of seven patients with nonobstructive azoospermia. Semen analysis confirmed that the adult survivor of childhood cancer was azoospermic. His cryopreserved testis cells were transplanted back into one testis, with no adverse effects. After transplantation, the testicular parenchyma had a normal homogeneous appearance. Testosterone, Follicle Stimulating Hormone, and Luteinizing Hormone levels were normal. Inhibin B levels were low. The patient remains azoospermic one year after transplantation.

**Interpretation:** We describe a platform for proficiency training in UGRT flushing, aspiration, or injection. This provides a noninvasive option for sperm retrieval and infusing stem cells or other therapeutics into the testis.

**Funding:** The *Eunice Kennedy Shriver* National Institute for Child Health and Human Development HD100197 and the UPMC Magee Center for Reproduction and Transplantation.

## INTRODUCTION

Azoospermia (no sperm in the ejaculate) can be caused by genetic mutations, chromosomal abnormalities, medical treatments (e.g., chemotherapy or radiation), or other circumstances. In some cases, sperm can be retrieved directly from the testes and used to achieve pregnancy with established assisted reproductive technologies. For adult patients with obstructive azoospermia (OA, blockage in the excurrent duct system), sperm can usually be retrieved directly from the testis or epididymis using testicular sperm extraction (TESE), testicular sperm aspiration (TESA), or percutaneous epididymal sperm aspiration (PESA). These procedures involve cutting or tearing seminiferous tubules or epididymal ductules from the testis or epididymis, respectively, which are teased apart in the andrology lab to release sperm. These blind approaches are less effective for patients with nonobstructive azoospermia (NOA) where rare patches of spermatogenesis, if present, are randomly dispersed in the testicular parenchyma.

Microsurgical testicular sperm extraction (microTESE) involves opening the testis to enable a detailed examination of all seminiferous tubules with a surgical microscope to identify rare tubules that are plump and may contain sperm. MicroTESE is invasive, time consuming, and expensive but leads to successful sperm retrieval in about 50% of NOA cases. Sperm from all seminiferous tubules are deposited in the rete testis space before exiting the testis. The rete testis is echodense and can be visualized on ultrasound. We will test the hypothesis that ultrasound-guided rete testis (UGRT) flushing and aspiration can be used to noninvasively retrieve sperm from the rete testis of men with azoospermia.

Chemotherapy and radiation treatments for cancer or other conditions can cause permanent infertility. Adult men have the option to cryopreserve sperm prior to treatment, which can be thawed in the future to achieve pregnancy using established assisted reproductive technologies.^1–3^ Those options are not available to prepubertal patients who are not yet producing mature sperm. Most children will survive their cancer and still have their entire reproductive lives in front of them.^4^ Adult survivors of childhood cancers desire to have children.^5–9^ Prepubertal males do have spermatogonial stem cells (SSCs) in their testes that have the potential to produce sperm. A method to transplant SSCs and regenerate spermatogenesis in chemotherapy-treated mice was pioneered by Brinster and colleagues in 1994^10,11^. SSC transplantation has been replicated in numerous animal species, leading to the production of donor-derived embryos or offspring in mice, rats, goats, sheep, and monkeys (reviewed in ^12^). In larger animals (e.g., goats, sheep, pigs, monkeys), UGRT injection is used to introduce SSCs into the seminiferous tubules of the testis. The rete testis space is contiguous with all seminiferous tubules where SSCs reside and where sperm are made. Therefore, an injection into the rete testis space can simultaneously fills all seminiferous tubules at the same time. We hypothesize that this method can be safely used for SSC transplantation in human patients.

Immature testicular tissues have been cryopreserved for over 3,000 patients worldwide in anticipation that SSCs in those tissues can produce sperm.^13^ Jensen and colleagues recently reported the autologous transplantation of cryopreserved testicular tissue in an adult man with idiopathic NOA.^14^ Here we report the first case of re-transplantation of cryopreserved immature testicular cells into an infertile adult survivor of childhood cancer using UGRT injection.

## METHODS

### Ultrasound-guided rete testis flushing and aspiration to retrieve sperm from the testes of monkeys with chemotherapy-induced azoospermia

As part of a separate study, the testes of six rhesus macaques were irradiated to eliminate endogenous spermatogenesis. Cryopreserved rhesus testicular cells from another monkey were then thawed and injected into the testes of the irradiated monkeys to regenerate spermatogenesis, as previously described.^15^ At the end of that study these monkeys were used as a platform to train the urologist (KH) in the technique of UGRT flushing and aspiration. A 25-gauge, 1·5-inch hypodermic needle was inserted under ultrasound guidance through the base of the scrotum until the tip emerged into the rete testis space. The rete testis was flushed with MEM-alpha containing trypan blue (10% v/v; Gibco, 15250061) and Optison (20% v/v; GE Healthcare, 2707-03) and then aspirated. Sperm in the aspirate were counted, centrifuged (1460 × g for 5 minutes), resuspended in 0·5 mL modified human tubal fluid (mHTF, Irvine Scientific, 90126) and 0·5 mL of TEST-yolk buffer containing glycerol and gentamicin (Irvine Scientific, 90128) and cryopreserved. Testes and epididymides were then harvested. Testes were cut into thick sections to confirm perfusion of blue dye from the rete testis space into the seminiferous tubules. The cauda epididymis was dissected to release and quantify sperm.

Sperm aspirated from the rete testis space were sent to the Assisted Reproductive Technology Core of the Oregon National Primate Research Center (ONPRC) to fertilize rhesus oocytes by intracytoplasmic sperm injection (ICSI), as previously described.^16^ Fertilization and pre-implantation embryo development were documented (Supplemental Materials). All monkey studies were reviewed and approved by the Institutional Animal Care and Use Committees of the MD Anderson Cancer Center and the Oregon Health and Science University.

### Ultrasound-guided rete testis flushing and aspiration to retrieve sperm from the testes of men with obstructive azoospermia or nonobstructive azoospermia

Ten men with azoospermia were enrolled in a study to retrieve sperm by UGRT flushing and aspiration (NCT03291522), as described for monkeys above. Approximately 500 µL of sterile 0·9% saline was flushed and aspirated from the rete testis space of both testes. Aspirates were then processed and analyzed for the presence of sperm. Three patients with OA chose to pair the experimental rete testis sperm aspiration procedure with a standard of care TESA procedure. Three of seven patients with NOA paired the experimental rete testis sperm aspiration procedure with the standard of care microTESE procedure. Sperm collected by experimental and standard of care approaches were quantified. All studies involving human subjects were reviewed and approved by the University of Pittsburgh Institutional Review Board.

### Testicular tissue biopsy, processing and cryopreservation

The UPMC Fertility Preservation Program and its coordinated centers in the US have performed testicular tissue cryopreservation (TTC) for 1,010 patients between January 2011 and January 2025 (NCT02972801, NCT05829928). Research patient TTC-003 was a peripubertal boy diagnosed with osteosarcoma of the femur and had a planned treatment regimen that was associated with a significant risk of infertility (Supplemental Methods). When the patient was in early stages of his cancer treatment, testicular tissue was collected by incisional biopsy comprising about 20% of one testis. Most of the tissue (75%) was processed to produce a testicular cell suspension (Supplemental Methods) that was cryopreserved for the patient’s future use. The remaining 25% of tissue was allocated to research.

### Autologous SSC transplantation in a human adult survivor of childhood cancer

Patient TTC-003 returned as a young adult (21-25 yo) survivor of childhood cancer. Two semen analyses indicated that he was azoospermic (no sperm in the ejaculate) and he was enrolled in a study to have his cryopreserved testicular cells transplanted back into one of his testes (NCT04452305). His oncologist and cardiologist provided medical clearance prior to the procedure. One vial of patient cells was removed from liquid nitrogen storage, thawed, and washed as detailed in Supplemental Methods. The resulting cell suspension was centrifuged and resuspended in 150 µL of injection medium (mHTF with 10% pulmozyme, 10% serum substitute supplement (SSS), and 20% Optison) and loaded into the transplantation apparatus. The rete testis was visualized by ultrasound (FUJIFILM SonoSite SII) and used to guide accurate insertion of a 25-gauge, 1.5-inch hypodermic injection needle into the rete testis space. Once positioned, cells (150 µL) were injected into the rete testis followed by approximately 300 µL of sterile PBS.

## RESULTS

### Ultrasound-guided rete testis flushing and aspiration to retrieve sperm (monkeys)

We devised a simple device for UGRT flushing and aspiration, featuring a 1cc syringe, connected via a 21-gauge luer stub adaptor to a 15 cm flexible polyethylene tube (inner diameter: 0.76 mm, outer diameter: 1.22 mm), which was connected via another luer stub adaptor to a male-male connector and a 25-gauge, 1.5-inch hypodermic needle (**Figure 1A**). The 25-gauge needle is small to minimize trauma but has the tensile strength to puncture and be accurately guided through the testicular parenchyma. The polyethylene tubing allows the user to have free range of motion to accurately insert and direct the needle. The rete testis and injection needle are echo-dense and can be visualized on ultrasound (**Figure 1B**). Ultrasound was used to visualize the echo-dense rete testis space and accurately guide the echo-dense injection needle through the base of the scrotum and into the rete testis space. UGRT flushing and aspiration resulted in successful sperm retrieval from four of the six monkeys. Sperm counts in the aspirates ranged from 666 to 76,000 and confirmed accurate targeting of the rete testis space. Testes and epididymides were then collected, and sperm were found in the cauda epididymis from five of the six monkeys. One monkey (13-092) was azoospermic, which explains why sperm were not successfully aspirated from the rete testis in that monkey (**Table 1**).^15^ Accurate targeting of the rete testis space was also confirmed by the presence of blue dye emanating from the rete testis into the seminiferous tubules in thick, whole mount sections of the injected testes (**Figure 1C**). Sperm that were retrieved from the rete testis space (**Figure 1D**) were tested functionally by ICSI at the Oregon National Primate Research Center, resulting in fertilization and pre-implantation embryo development to the morula stage (**Figure 1E**).

**Table 1:** Sperm retrieval from rhesus macaque monkeys by UGRT flushing and aspiration and dissection of the cauda epididymis.

|  | Age (Yr) | Diagnosis | Additional Treatment | Sperm found with Rete Aspiration | Sperm found in Cauda Epididymis (x10<sup>6</sup>) |
| --- | --- | --- | --- | --- | --- |
| Monkey 13-092 | 3·4 | NOA Post-radiation | SSC Transplantation | 0 | 0 |
| Monkey 13-094 | 3·4 | NOA Post-radiation | SSC Transplantation | 60,000 | 4·7 |
| Monkey 13-114 | 3·5 | NOA Post-radiation | SSC Transplantation | 5,284 | 26 |
| Monkey 13-120 | 3·4 | NOA Post-radiation | SSC Transplantation | 666 | 0·6 |
| Monkey 13-122 | 3·5 | NOA Post-radiation | SSC Transplantation | 0 | 0·9 |
| Monkey 13-124 | 3·4 | NOA Post-radiation | SSC Transplantation | 76,000 | 13 |

Abbreviations: NOA, nonobstructive azoospermia

**Figure 1.**
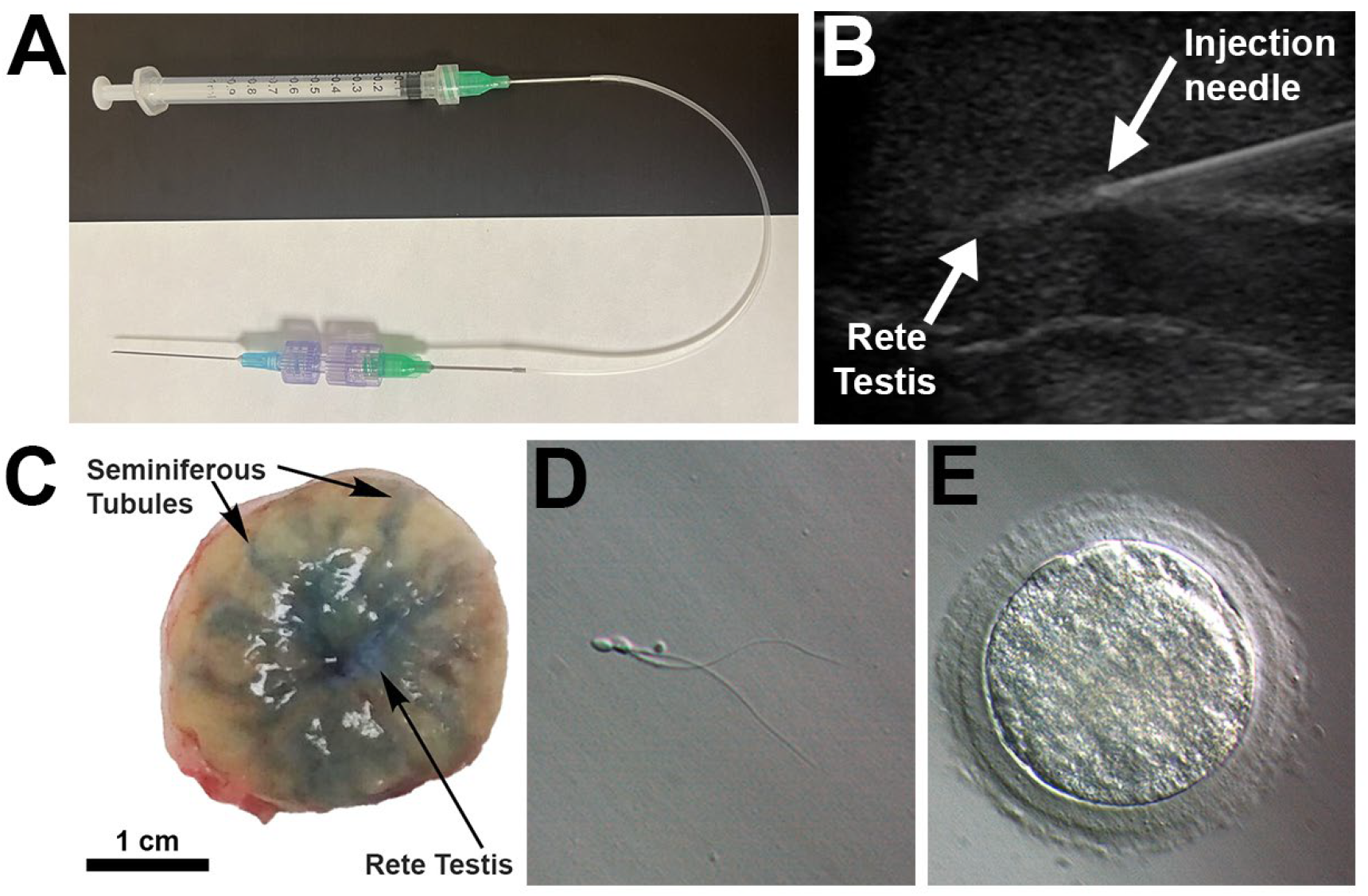
Ultrasound-guided rete testis flushing and aspiration in monkeys. A simple rete testis injection/aspiration device features a 1 cc syringe connected via luer stub adaptors to a flexible polyethylene tube, a male-male connector and a 25-gauge, 1.5-inch hypodermic needle (Panel A). The echo-dense rete testis space and injection needle are visible on ultrasound (Panel B). Thick, whole mount sections of testis reveal that blue dye emanated from the rete testis space into the distal seminiferous tubules indicating accurate targeting of the rete testis (Panel C). Normal testicular sperm aspirated from the rete testis (Panel D) were competent to fertilize rhesus oocytes, leading to preimplantation embryo development to the morula stage (Panel E).

### Ultrasound-guided rete testis flushing and aspiration to retrieve sperm (patients)

UGRT flushing and aspiration resulted in successful sperm recovery in three out of three OA patients (3, 119, and 360 sperm, respectively) and zero out of seven NOA patients. The experimental UGRT flushing and aspiration procedure was followed by the standard of care TESA procedure in all three OA patients and microTESE in three of the seven NOA patients. Sperm were successfully recovered by TESA from all three OA patients and by microTESE from two out of three NOA Patients (**Table 2**). No adverse events were reported for any of the azoospermia patients related to the UGRT flushing and aspiration procedure.

**Table 2:** Sperm retrieval from patients by UGRT flushing and aspiration and standard of care sperm retrieval techniques.

|  | Age (Yr) | Diagnosis | Sperm retrieved by rete testis aspiration | SOC Procedure | Sperm retrieved by SOC procedure |
| --- | --- | --- | --- | --- | --- |
| Patient 1 | 31-35 | NOA | 0 | MicroTESE | 0 |
| Patient 2 | 31-35 | NOA | 0 | MicroTESE | 2 |
| Patient 3 | 26-30 | NOA | 0 | NA | NA |
| Patient 4 | 26-30 | NOA | 0 | MicroTESE | 1 |
| Patient 5 | 36-40 | OA | 119 | TESA | 630,000 |
| Patient 6 | 36-40 | OA | 360 | TESA | 935,000 |
| Patient 7 | 31-35 | OA | 3 per HPF | TESA | 41 per HPF |
| Patient 8 | 31-35 | NOA | 0 | NA | NA |
| Patient 9 | 31-35 | NOA | 0 | NA | NA |
| Patient 10 | 41-45 | NOA | 0 | NA | NA |

Abbreviations: SOC, standard of care; NOA, nonobstructive azoospermia; MicroTESE, microsurgical testicular sperm extraction; NA, not applicable; OA, obstructive azoospermia; TESA; testicular sperm aspiration; HPF, high power field.

### Testicular biopsy and cell suspension

Pathological assessment of the biopsied testicular tissue from TTC-003 confirmed normal testis morphology and no malignancy (**Figure 2A**). Immunofluorescence staining of the research portion of the patient tissue confirmed the presence of VASA+ germ cells (2·14 cells/tubule) and UTF1+ stem/progenitor spermatogonia (1·86 cells/tubule) in the seminiferous tubules (**Figure 2B, C, and D**), even though this patient was already in the early stages of his treatment. The remaining tissue was digested with enzymes to produce a cell suspension (25·58 × 10^6^ cells; 94% viable). Two vials were cryopreserved for the patient’s future use.

**Figure 2.**
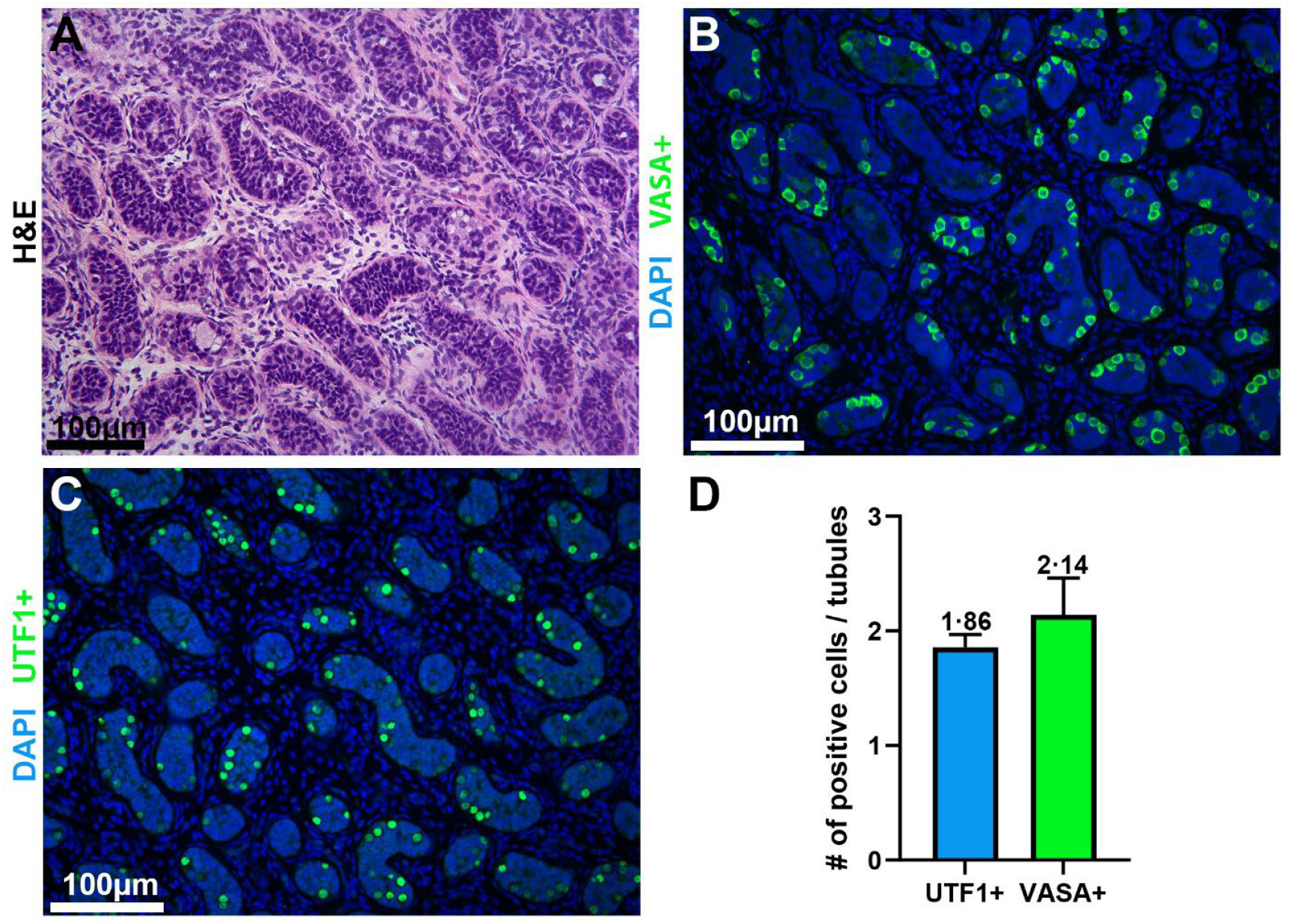
Histological and molecular analysis of patient testicular tissue. Histological section (5 µm) of patient testicular tissue biopsy counterstained with hematoxylin and eosin (H&E) showed normal immature testis morphology (Panel A). Immunohistochemistry (IHC) analysis revealed the presence of VASA+ germ cells (green) (Panel B). IHC revealed the presence of UTF1+ stem/progenitor spermatogonia (green) (Panel C). DAPI (blue) labels all cell nuclei (Panels B and C). Quantification of UTF1+ and VASA+ cells per seminiferous tubule (Panel D).

### Ultrasound-guided rete testis injection of cryopreserved autologous prepubertal testicular cell suspension

The peripubertal osteosarcoma patient returned as a young adult, to have his cryopreserved testicular cells transplanted back into his testis. Ultrasound indicated that his testes were small (right testicle: 9·42 mL; left testicle: 9·6 mL (normal range 17-50 mL)). Two semen analyses revealed no sperm in the ejaculate (azoospermia). Testosterone (442 ng/dL), FSH (12·4 mIU/mL), and LH (4·5 mIU/mL) were within normal ranges. Inhibin B (32 pg/dL) was below the normal range (**Table 3**).

**Table 3:** Clinical testing before and after autologous testis cell transplantation.

|  | Pre-Tx Values | 6-month Post-Tx Values | 1-year Post-Tx Values | Normal Range | Comments |
| --- | --- | --- | --- | --- | --- |
| **Total Testosterone** | 442 ng/dL | 439 ng/dL | 375 ng/dL | 250 - 1,100 ng/dL | Normal |
| **Free Testosterone** | No data | 83 pg/mL | 91·1 pg/mL | 35 - 155 ng/dL | Normal |
| **Follicle Stimulating Hormone (FSH)** | 12·4 mIU/mL | 11·3 mIU/mL | 10·7 mIU/mL | 1·5 - 12·4 mIU/mL | Normal/high |
| **Luteinizing Hormone (LH)** | 4·5 mIU/mL | 12·6 mIU/mL | 5·2 mIU/mL | 1·7 - 8·6 mIU/mL | Normal/high |
| **Inhibin B** | 32 pg/dL | 18 mpg/dL | 30 pg/mL | 47 - 308 pg/dL | Low |
| **Right testis volume (Ultrasound)** | 9·42 mL | 10 mL | 9·14 mL | 17 - 50 mL | Small, normal parenchyma |
| **Left testis volume (Ultrasound)** | 9·6 mL | 9·24 mL | 8·27 mL | 17 - 50 mL | Small, normal parenchyma |
| **Sperm count** | 0 | 0 | 0 | 15 - 200 million/mL | Azoospermic |
| **Semen volume** | 4·3 | 2·9 mL | 3·7 mL | > 1·1 mL | Normal |
| **Semen pH** | 8 | 7·6 | 8 | ≥ 7·2 | Normal |

Abbreviations: Tx, Transplantation; mIU, milli international units

One vial of patient testicular cells was thawed, yielding 9·4 × 10^6^ viable cells (12·9 × 10^6^ cells; 72·9% viable), which were washed and resuspended in 150 µL of injection medium. The transplantation apparatus was primed by loading 800 µL of sterile PBS followed by 150 µL testicular cell suspension with a small air gap in front of and behind the cell suspension (**Figure 3A**). Air bubbles in front of the cell suspension can be visualized by ultrasound and used to confirm that the needle is correctly positioned when bubbles fill the rete testis and migrate into the seminiferous tubules (**Figure 3B-D**). The patient reported being pain free within a few days after transplantation and did not experience any adverse effects.

**Figure 3.**
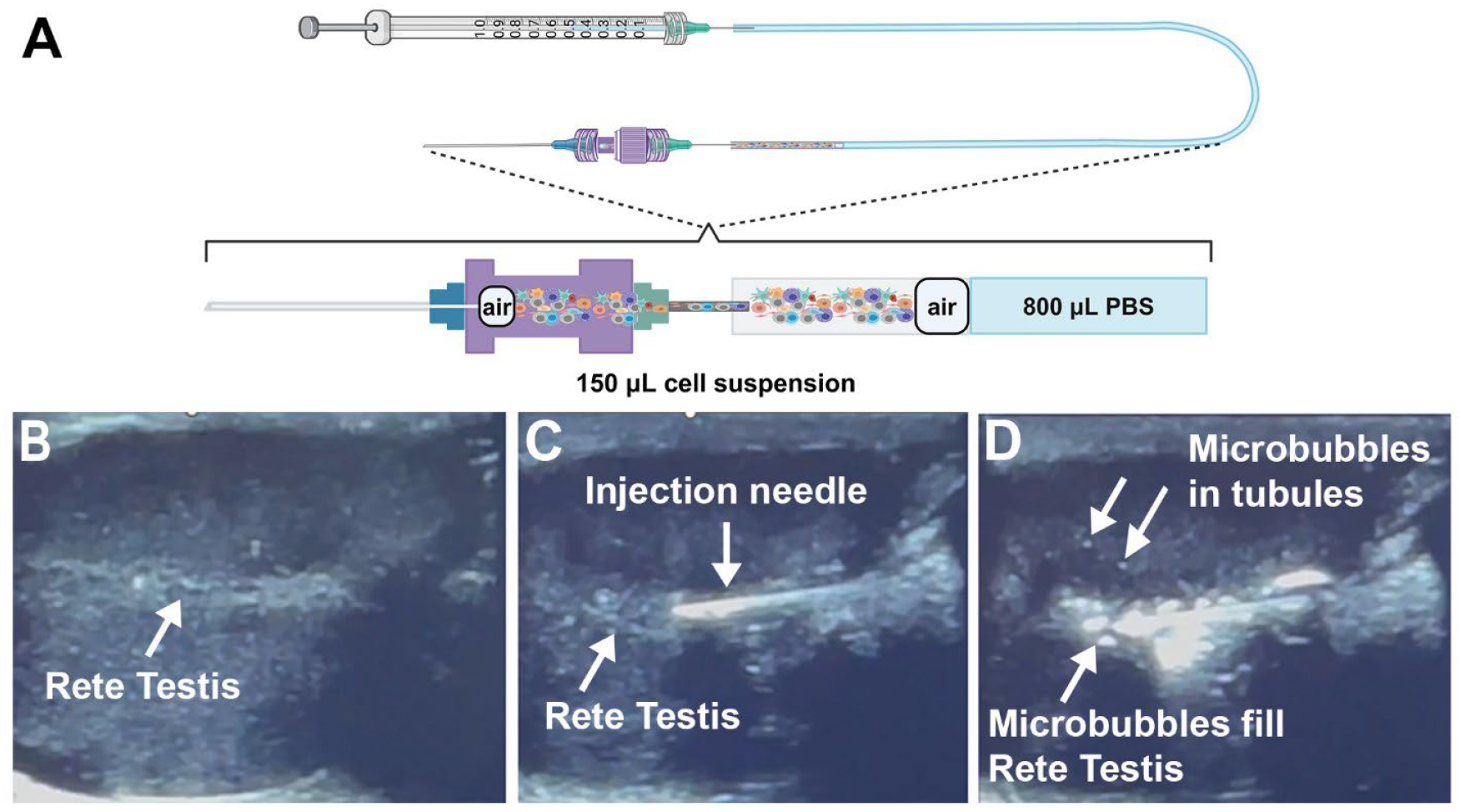
Ultrasound-guided rete testis injection of cryopreserved testicular cells in an adult survivor of childhood cancer. The UGRT injection device was loaded with PBS (800 µL), followed by a small air bubble, 150 µL of the patient cell suspension, and another small air bubble (Panel A). The rete testis was visualized by ultrasound (Panel B). The injection needle was inserted through the base of the scrotum and advanced until the needle emerged into the center of the rete testis space (Panel C). The air bubble in front of the patient cell suspension filled the rete testis space with bubbles that migrated into the seminiferous tubules (Panel D).

Six-month and 12-month follow-up ultrasound assessments indicated that both testes remained small with a normal homogeneous parenchyma. Testosterone, FSH, and LH levels were in the normal range. Inhibin B was below normal range. Semen volume and pH were normal. No sperm were found in the ejaculate (azoospermic) (**Table 3**).

## DISCUSSION

We were fortunate to have access to monkeys that served as a training platform for UGRT injection/aspiration, but this platform is not broadly accessible. On the other hand, fertility clinics frequently perform testicular sperm retrieval techniques such as TESA or microTESE in patients with azoospermia. This offers an opportunity to practice accessing the rete testis space under ultrasound guidance and poses minimal additional risk to the patient who is undergoing a more invasive surgical procedure to retrieve sperm. Furthermore, it may offer a less invasive, less expensive approach to sperm aspiration. Our results indicate that UGRT flushing and aspiration is effective for sperm retrieval in patients with OA but perhaps not NOA where the extent of spermatogenesis in the testis is expected to be limited. UGRT flushing and aspiration is not surgical, so is less painful, less time-consuming and will likely be less expensive than standard of care sperm retrieval options. Indeed, several of our patients opted to forego the standard of care sperm retrieval procedure if sperm were retrieved using the UGRT approach.

Radford and colleagues reported the first autologous transplantation of testicular cells that had been cryopreserved for 12 adult patients with Hodgkin’s disease. Seven of those patients returned to have cells transplanted back into their testes, via the rete testis. The method for targeting the rete testis in that study was not described but a cited reference^17^ suggests that ultrasound may have been used. The outcomes of that study were not reported.^18,19^

This is the first report of re-transplantation of cryopreserved immature testicular cells into the testes of an adult survivor of childhood cancer. TTC-003 remains azoospermic and we will continue to monitor testicular volume and structure by ultrasound, semen analyses, and hormones on a biannual basis. If sperm are observed in the ejaculate, they can be used immediately to fertilize partner oocytes or cryopreserved for future use. If no sperm are observed in the ejaculate and the patient is ready to start his family, he can attempt to retrieve sperm from the testis by microTESE.

It is important to manage expectations of the patients, their families and the medical/research community. The size of biopsy collected when our young patient (~20% of one testis) was small and probably contained a small number of stem cells. It is reasonable to expect that those cells will engraft and produce pockets of spermatogenesis in the testis, as observed in several animal models.^12^ However, the extent of spermatogenesis may be limited and not be sufficient for any sperm to emerge in the ejaculate. Methods to maintain and expand human SSCs in culture remain elusive (reviewed in^20^) but if successful, could increase the number and regenerative potential of SSCs in the biopsies from young patients.

Our study developed a non-invasive approach for accessing the rete testis space in patients. We devised a simple injection apparatus and established a training platform that is accessible to experts in men’s reproductive health and their trainees. Finally, we demonstrated the safety and feasibility of accessing the rete testis space to aspirate sperm or infuse autologous stem cells into the testes.

## Data Availability

All data produced in the present study are available upon reasonable request to the authors

## AUTHOR CONTRIBUTIONS

SLD, GMC, DAG, KH, and KEO counseled and consented study participants. GMC recovered testicular biopsy from prepubertal patient, TTC-003. KAP processed and cryopreserved testicular tissues for TTC- 003. ACZ thawed and prepared TTC-003 samples for transplantation. GS, MLM, KH, and KEO performed rete testis aspirations in monkeys. CBH and team at the Oregon National Primate Research Center performed ICSI with sperm aspirated from monkey rete testes. MH performed fertility assessments on TTC-003 and referred to the study. AW, KH, KEO, and team did ultrasound guided rete testis aspiration and/or injection of cells. KAP, ACZ, GS, MLM, and CBH analyzed patient and monkey data. ACZ and KEO wrote the manuscript.

## ACKNOWLEDGEMENTS

We thank the Assisted Reproductive Technology Core at the Oregon National Primate Research Center for help with the monkey ICSI experiments. The lab animal research team at MD Anderson Cancer Center helped with the monkey rete testis aspiration experiments. The UPMC Magee Center for Reproduction and Transplantation provided the infrastructure and funding for rete testis aspiration and SSC transplantation procedures in patients.

## Supplementary Appendix

This appendix has been provided by the authors to give readers additional information about their work.

Supplementary Appendix to:

### METHODS

#### Oncologic treatment for patient

Patient TTC-003 had already initiated treatment at the time of testicular tissue cryopreservation with a prior exposure to gonadotoxic agents 2.8 g/m^2^ ifosfamide (CED = 0.7 g/m^2^), 180 mg/m^2^ cisplatin, as well as 60 mg/m^2^ etoposide, 168.75 mg/m^2^ doxorubicin, and 72 g/m^2^ methotrexate. The patient’s overall treatment regimen for osteosarcoma of the femur followed a clinical oncology protocol AOST0331, which included a cumulative total of 14.4 g/m^2^ ifosfamide (cyclophosphamide equivalent dose (CED^1^= 3.5 g/m^2^) and 240 mg/m^2^ cisplatin that are gonadotoxic; in addition, the protocol includes 300 mg/m^2^ etoposide, 144 g/m^2^ methotrexate, and 225 mg/m^2^ doxorubicin.

#### Testicular tissue biopsy, processing and cryopreservation

Testicular tissue was cut into small pieces that were teased apart with sterile forceps and then digested with 1mL of Liberase MTF C/T (Roche, 05339880001; 2mg (8U)/mL) with 60µg/mL Thermolysin in 1X PBS per 100mg of tissue for 8 minutes at 37°C with agitation (250 rpm). Digested tissue was centrifuged 200 × g for 5 minutes, the supernatant discarded, and then washed with fresh 1x PBS twice before digestion with 0.25mL Pulmozyme (Genentech; 1mg/mL) and 1mL of Trypzean (Sigma Aldrich, T3568; 2.5mg/mL) in PBS with 1.4mM EDTA per 100mg of tissue for 15 minutes at 37°C in a water bath. Digestion was terminated by adding 10% volume of Serum Substitute Supplement (SSS; Irvine Scientific, 99193) and the suspension was filtered through a 70µm cell strainer. Cells were pelleted by centrifugation at 600 × g for 15 minutes and resuspended in 15% SSS 10%DMSO (Gaylord and Chemical, Procipient®) 75% Quinn’s Advantage Blast Medium (Cooper Surgical, ART 1029) at a cell concentration of 1×10^7^ cells/mL. Cells were frozen in a Planer controlled-rate freezing machine (−1°C/minute to − 40°C followed by −10°C/minute to −90°C).

#### Functional testing of monkey sperm retrieved by ultrasound-guided rete testis flushing and aspiration: Fertilization and preimplantation embryo development

Rhesus oocytes were obtained by controlled ovarian stimulation of female rhesus macaques, as previously described.^2,3^ The resulting oocytes were fertilized by intracytoplasmic sperm injection (ICSI)^4–6^ using sperm that were retrieved by ultrasound-guided rete testis flushing and aspiration. After ICSI, injected oocytes were placed in 4-well dishes (Nalgene-Nunc) containing 500 µl of BO-IVC medium (IVG Bioscience), covered with light mineral oil and cultured at 37°C in 6% CO_2_, 5% O_2_ and 89% N_2_. Embryos were cultured for 8 days or until embryo development arrested.

#### Thawing and preparing cells for autologous SSC transplantation in a human adult survivor of childhood cancer

One vial of patient cells was removed from liquid nitrogen storage and quickly thawed in a 37°C water bath (less than 3 minutes). The thawed cell suspension was transferred to a 15 mL conical tube and washed twice with modified Human Tubal Fluid (mHTF; Irvine Scientific, 90126) containing 10% SSS. Cells were centrifuged at 600 × g for 5 minutes. The supernatant was removed, and cells were resuspended in mHTF with 10% SSS and 0.1% pulmozyme (1mg/mL). At this step, 2µl of cell suspension was removed, diluted 1:1 with trypan blue, and used to calculate cell number and viability. Cells were centrifuged at 600 × g for 5 minutes. Supernatant was carefully removed, 15 µl of pulmozyme (1mg/mL) was added and the tube was flicked to dislodge and disaggregate the pellet. Additional injection medium sans pulmozyme was added to make the final volume 150µl of cell suspension in mHTF with 10% pulmozyme, 10% SSS and 20% Optison which was loaded into the transplantation apparatus.

